# Promoting early childhood development through nurturing parenting: Feasibility of the ASCENDI program for adolescent mothers in the Dominican Republic

**DOI:** 10.64898/2026.09.15.26362692

**Authors:** Nora Badoui-Rodríguez, Lourdes Pérez, Hilcemery Fortuna-Mejía, Laura V. Sánchez-Vincitore, Arachu Castro

## Abstract

This study evaluated the feasibility and preliminary effectiveness of a 12-session group-based nurturing care intervention for adolescent mothers in the Dominican Republic. We conducted a one-group pretest-posttest feasibility study in Boca Chica, Dominican Republic. Participants were adolescent mothers under 20 years of age with a child enrolled in an early childhood development (ECD) center operated by the National Institute for Early Childhood Comprehensive Care. The intervention, based on the World Health Organization’s Nurturing Care Framework, addressed ECD, affective care, positive discipline, and maternal well-being. We calculated effect sizes using Cohen’s *d*.

Of 14 adolescent mothers who started the program, 11 completed both assessments (78.6% retention). Participants had a mean age of 16.91 years (*SD* = 1.35), with children averaging 1.15 years (*SD* = 0.84). Two outcomes showed statistically significant improvements. Parental efficacy (1–4 scale) increased from *M* = 3.38 to *M* = 3.84 (*d* = 0.99, *p* = .008), with significant gains observed in three subdomains: protective (*d* = 1.21, *p* = .003), formative (*d* = 0.90, *p* = .014), and bonding competencies (*d* = 0.81, *p* = .023). Cognitive stimulation activities (0–1 scale) increased from *M* = 0.42 to *M* = 0.65 (*d* = 0.81, *p* = .036). Participant satisfaction was high (*M* = 4.64 on a 5-point scale).

ASCENDI demonstrated feasibility and preliminary effectiveness in improving parental efficacy and cognitive stimulation among adolescent mothers. Large effect sizes and high retention and satisfaction support the potential of group-based nurturing care interventions for adolescent mothers in resource-limited settings.

## Introduction

Adolescent mothers face compounding adversities that affect both their well-being and their children’s development. They are disproportionately exposed to socioeconomic disadvantage, limited educational opportunities, gender-based violence, and psychological distress, all of which can impact their parenting abilities (Kofke et al., 2022; Nelson, Frías, et al., 2025; Nelson, Sánchez-Vincitore, et al., 2025; Nguyen et al., 2019; Sánchez-Vincitore & Castro, 2022). Some studies have found that, compared to older mothers, adolescent mothers have fewer skills in self-regulation, stress management, patience, and understanding their children’s needs, and that these differences may result in less responsive, stimulating, and nurturing care and in greater use of physical and psychological violence (Britto et al., 2017; Buzi et al., 2020; Hubert et al., 2021; Mwenda et al., 2023). However, these differences are not universal: across 15 Latin American and Caribbean countries, adolescent mothers used significantly more violent discipline than older mothers only in Honduras after adjustment for socioeconomic covariates (Castro et al., 2026). Adolescent mothers also have fewer material resources for stimulation, such as books and toys, whose absence is associated with early childhood development (ECD) delay (Chang et al., 2024). Their children, in turn, face elevated risks of adverse health outcomes, including low birth weight, preterm birth, and undernutrition; perinatal, neonatal, post-neonatal, and child mortality; poor physical and mental health; ECD delay; and lower language development, educational attainment, life satisfaction, and personal income (Falster et al., 2018; González-Rodríguez et al., 2024; Kumar & Huang, 2021; Nguyen et al., 2019; SmithBattle et al., 2024; Welch et al., 2024).

Secondary data analysis of the 2018 Indonesian National Socioeconomic Survey and Basic Health Survey reported that up to 60% of adolescent mothers did not provide comprehensive care, with their children at greater risk of developmental delay (Sumiati et al., 2023). Similarly, a 2015 prospective cohort study conducted in Brazil, Guatemala, India, the Philippines, and South Africa found that children of mothers younger than 20 had lower secondary education completion rates (Fall et al., 2015). These challenges are not solely attributable to the mother’s age: whereas some scholars have argued that poverty is the most important explanatory variable of ECD delay among children of adolescent mothers (Geronimus et al., 1994), others have emphasized that parenting practices compounded with preexisting and ongoing social disadvantages help explain the association between adolescent motherhood and lower ECD outcomes (Mollborn & Dennis, 2012). Research using the life course approach has found that the social disadvantages that contribute to adolescent pregnancy are transmitted intergenerationally, and that having resources such as income or maternal education counteracts the adverse effects of adolescent pregnancy (Mollborn & Dennis, 2012). A cross-national analysis of 15 Latin American and Caribbean countries found that socioeconomic disadvantage accounted for most of the ECD gap among children of adolescent mothers (Castro et al., 2026). In that analysis of 23,650 children aged 36–59 months, children of adolescent mothers had lower ECD scores in 11 of the 15 countries, but after adjustment for household wealth, maternal education, and urban or rural residence, the association persisted only in Paraguay and Suriname. Understanding and addressing the adversities that adolescent mothers themselves experience is therefore essential to improving outcomes for both mothers and children (Castro & Sánchez-Vincitore, 2026a).

Adolescent pregnancy is prevalent in the Dominican Republic, where an estimated 25,120 adolescents (19.1% of live births) gave birth in 2023 (United Nations, 2024). An analysis of 2014 and 2019 Multiple Indicator Cluster Survey (MICS) data showed that violent discipline was associated with detrimental effects on child development, that children of adolescent mothers had significantly lower development scores than children of mothers aged 20 and above even after controlling for children’s ages, and that mothers who rejected physical punishment provided more stimulation activities even without physical resources such as books or toys (Sánchez-Vincitore & Castro, 2022). That analysis also found that mothers who believed in physical punishment implemented both violent and positive disciplinary strategies, demonstrating that these are not mutually exclusive and that we should not assume that the presence of the latter implies better practices (Sánchez-Vincitore & Castro, 2022). However, in a 2024–2025 study with 1,019 mothers participating in free, public early childhood programs provided by the National Institute for Early Childhood Comprehensive Care (INAIPI), we found that violent discipline increased with maternal age at first childbirth (*r* = 0.159, *p* < .001) and that former adolescent mothers used significantly less violent discipline than adult-onset mothers (*d* = −0.22, *p* < .001), a difference that was robust to covariate adjustment and that, if replicated, would challenge deficit perspectives and support strengths-based approaches (Castro & Sánchez-Vincitore, 2026a). In the 15-country MICS analysis, which included the 2019 Dominican Republic survey, the adjusted difference in violent discipline between adolescent and adult mothers in the Dominican Republic was negligible (β = −0.004), and adolescent mothers used more positive discipline (β = 0.052, *p* = .024) (Castro et al., 2026). Neither dataset, therefore, supports the view that adolescent mothers in the Dominican Republic use more violent discipline than older mothers.

The provision of nurturing care has favorable effects on attachment, child growth, and exclusive breastfeeding, with the latter having positive impacts, reducing child morbidity and mortality and significantly increasing adolescent intelligence and educational achievement (Bar et al., 2016; Brahm & Valdés, 2017; Britto et al., 2017; Couto et al., 2020; Herba et al., 2013). In the Dominican Republic, a 2023 study with a sample of 699 children aged 12–36 months found that ever-breastfed children had higher scores in overall ECD than those who were not; higher scores in language and fine motor development primarily drove this effect, and the never-breastfed group had a greater risk of developmental delay in fine motor and socioemotional development (Sánchez-Vincitore et al., 2024).

A safe, responsive, and nurturing caregiving environment supports adequate emotional and cognitive development and mitigates the adverse effects of growing up in vulnerable contexts (Britto et al., 2017). The capacity to provide such care is related to how mothers themselves received care, with adverse experiences presenting a high risk of generating a cycle of maltreatment (Britto et al., 2017; Nelson, Frías, et al., 2025). Child maltreatment is associated with adverse child development, behavioral and emotional problems, and poor school performance (Petersen et al., 2014). Yet as of 2025, less than half of children globally grew up in home environments that provide early stimulation and responsive care (UNICEF, 2024), and between 2010 and 2016 an estimated more than three-fifths of children in low- and middle-income countries were subjected to physical aggression (Cuartas et al., 2019).

Prevention programs promoting nurturing care offer a promising approach to preventing violent parenting and interrupting the intergenerational transmission of trauma. There is evidence that children of mothers with an adequate level of maternal health literacy—cognitive and social skills that determine the motivation and ability of women to understand and use information in ways that promote and maintain their own and their children’s health—have better scores on development scales (Oflu & Yalcin, 2023). Interventions aimed at fostering nurturing and responsive parenting have a positive effect on maternal health and on the development and health of their children, including increased maternal self-esteem, knowledge of appropriate caregiving practices, and enhancements in children’s motor, cognitive, and socioemotional development (Fisher et al., 2023; McCoy et al., 2020; Tofail et al., 2023). A systematic review of 21 randomized clinical trials in low- and middle-income countries found that 12-session group programs were the most effective in improving ECD, with combined interventions addressing both nutrition and child development achieving a higher level of improvement and with no significant additional effects observed beyond 12 sessions (Zhang et al., 2021). Effects were greater among vulnerable populations due to a catch-up phenomenon whereby children and caregivers experiencing multiple inequalities showed accelerated developmental gains when provided with appropriate support (Zhang et al., 2021). Another systematic review of 56 studies across 22 countries found that parenting interventions can prevent physical and psychological violence against children (*d* = −0.46; 95% CI: −0.59, −0.33), with effects sustained up to 24 months (Backhaus et al., 2023). Similarly, a Malaysian parenting program developed between November 2018 and April 2019, which focused on parental roles, goals, and responsibilities, positive relationships and quality time, communication and positive reinforcement, health and safety through routines and rules, and effective discipline and conflict management, found reductions in global child maltreatment scales, physical abuse, emotional abuse, propensity toward physical punishment, feelings of parental inefficacy, and child behavioral problems (Lachman et al., 2023).

The World Health Organization (WHO) strongly recommends that all children between 0 and 3 years old receive responsive care and engage in early learning activities, with adequate support for parents and caregivers in environments that facilitate responsive parenting and early learning (WHO, 2023). However, existing parenting programs have been targeted at parents, mothers, and caregivers of any age without differentiated interventions for adolescent mothers living in adversity (Sumiati et al., 2023), despite evidence that adolescent mothers face distinct challenges requiring tailored support (Wuermli et al., 2021). There is nonetheless evidence of the benefits of interventions to improve adolescent mothers’ knowledge of care and parenting: a study conducted in Kenya found that an intervention based on a messaging platform and participation in a psychosocial support group was associated with improvements in global child development and in fine and gross motor development (Mwenda et al., 2023).

Including children’s fathers and promoting support networks are also relevant to nurturing care. Children of absent fathers have lower neurological and socioemotional development (Chang et al., 2024), whereas children whose fathers are present but implement negative parenting and disciplinary practices also have worse health outcomes (Heward-Belle & O’Leary, 2023). Promoting father participation from the prenatal stage is associated with stronger bonds with children during school age and adolescence (Fagan et al., 2023), improved overall child development (Yogman & Eppel, 2022), and improved mothers’ mental health (Kumar & Huang, 2021). Participation may not occur spontaneously; the father’s self-efficacy affects his involvement in play and parenting, making it important to encourage the paternal role and the social support that facilitates the acquisition of parenting and play skills (Bowles et al., 2022; Yoo, 2022).

Existing parenting programs differ in their objectives and implementation strategies. The evidence in the systematic review of parenting interventions in low- and middle-income countries was insufficient to determine the superiority of one implementation setting over another (e.g., home-based vs. group-based), though each had different advantages; in the case of group-based interventions, the review highlighted their broader reach, lower cost, and ability to facilitate peer support (Zhang et al., 2021). Parenting interventions integrated with other intersectoral programs promoting health, child nutrition, child protection, and responsive caregiving yielded better results (Britto et al., 2017). Table 1 describes five of these programs; none was designed specifically for adolescent mothers living in contexts of adversity.

**Table 1.** Core parenting intervention programs.

| Name of program | Program description |
| --- | --- |
| Reach Up and Learn Early Childhood Parenting Program (Caribbean Institute for Health Research, 2021) | A program for low-resource settings based on the Jamaica Home Visit intervention by Sally Grantham-McGregor, adapted in Bangladesh and Colombia, demonstrating benefits for child development and implemented on a large scale by the Peruvian government. |
| Programa Nacional Wawa Wasi (PNWW) (Hartinger SM et al., 2017) | A program from the Ministry of Women and Social Development (MIMDES) of Peru that provides comprehensive early childhood care aiming to promote children's cognitive, social, motor, and intellectual development, as well as their autonomy and independence. |
| SPARK Center at Boston Medical Center (Boston Medical Center, n.d.) | A center offering therapeutic and specialized programs for children with complex medical conditions and emotional and behavioral problems, as well as those affected by family hardships such as abuse or neglect. |
| The SHINE trial (Gladstone MJ et al., 2019) | A program focused on early nutrition and interventions to improve access to water, sanitation, and hygiene for children with and without HIV exposure. |
| Breastfeeding counseling (UNICEF/WHO) (Ara et al., 2019) | A course promoting exclusive breastfeeding. |

Given this gap, we designed the Adolescent Maternity: Support for Nonviolent Discipline and Early Child Development Initiative (ASCENDI) program in collaboration with INAIPI, which provides free early childhood services to approximately 212,500 children from 45 days to five years in areas of high social vulnerability. Until this study, INAIPI had not offered nurturing care programs specific to adolescent mothers. The intervention was based on WHO’s Nurturing Care Framework (WHO, 2018) and WHO’s guidelines on parenting interventions (WHO, 2020, 2023), and informed by our previous community-based research (Castro & Sánchez-Vincitore, 2025, 2026a, 2026b; Nelson, Frías, et al., 2025; Nelson, Sánchez-Vincitore, et al., 2025). The WHO guidelines emphasize adapting interventions to the local context, including nonviolent discipline techniques, positive reinforcement, parental self-regulation strategies, strategies to strengthen the parent-child relationship, mental health support guidance for caregivers, and psychosocial guidance to provide mothers with coping and social support (WHO, 2023). The objective of this study was to evaluate the feasibility and preliminary effectiveness of ASCENDI in improving parenting practices and cognitive stimulation activities among adolescent mothers.

## Materials and methods

### Participants

In collaboration with INAIPI, we surveyed two communities in greater Santo Domingo with the highest percentage of adolescent mothers to assess interest and availability. We selected the INAIPI network in Boca Chica, a community on the Caribbean coast approximately 32 km (20 miles) east of the capital city of Santo Domingo. Inclusion criteria were being a mother under 20 years of age with a child enrolled in an INAIPI center in Boca Chica. Exclusion criteria were severe mental health or cognitive conditions preventing participation, or concurrent participation in a similar parenting intervention.

### Ethics statement

The Institutional Review Boards of Tulane University (study 2025-054) and Universidad Iberoamericana (study CEI2025-42) approved the study on August 6, 2025, and February 10, 2025, respectively. We obtained informed assent from participants under 18 and informed consent from their parent or legal guardian in paper format; one participant aged 19 provided her informed consent. The privacy rights of study participants have been observed.

### Intervention

The ASCENDI intervention comprises 12 weekly group sessions organized in four modules: (1) Care and Caregiving, addressing child development, affective care, basic psychosocial skills, stress management and caregiver self-care, positive discipline, play and early learning, and nutrition and health; (2) Acknowledging Lived Experience, Emotions, and Resources, covering emotional regulation and communication, social support and community resources, and healing from adverse childhood experiences (ACEs); (3) Planning for the Future, focused on life projects toward self-sufficiency; and (4) Reflecting on Changes in Parenting (Table 2).

**Table 2.** Nurturing care for adolescent mothers: session topics, objectives, content, and WHO domains.

| Module | Session | Topic | Title | Objective | Content | Evidence-based recommendation (WHO, 2020, 2023) | Nurturing care framework component (WHO, 2018) |
| --- | --- | --- | --- | --- | --- | --- | --- |
| Care and Caregiving | 1 | Early Childhood Development (0-5 years) | Can you introduce me to your child? | Strengthen the bond between mother and child and understand early childhood development | Affective bond, developmental milestones, play, and communication | • Responsive caregiving<br>• Promote early learning | • Good health<br>• Responsive caregiving<br>• Opportunities for early learning |
|  | 2 | Affective Care | This is how I take care of my child | Reflect on care routines and their relationship with the expression of affection and children's needs | Developmental areas, children's needs and rights, healthy habits, and routines | • Responsive caregiving<br>• Promote early learning | • Good health<br>• Responsive caregiving |
|  | 3 | Basic Psychosocial Skills | My child under 5 years old as a person to care for and love | Understand psychosocial development in children and its relationship with caregivers | Basic trust, autonomy, initiative, and positive relationships | • Promote early learning | • Good health<br>• Responsive caregiving<br>• Opportunities for early learning |
|  | 4 | Stress Management and Caregiver Self-Care | Everyday Pressure and Stress | Recognize and manage stress associated with caring for young children | Parental stress, support networks, and stress reduction strategies | Supporting maternal mental health | Good health<br>Security and safety |
|  | 5 | Positive Discipline in Early Childhood | Tantrums and Outbursts: Disciplining Positively | Question discipline patterns and promote positive regulation in children | Positive discipline, respectful limits, and avoiding violence | Responsive caregiving | Good health<br>Security and safety<br>Opportunities for early learning |
|  | 6 | Play and Early Learning | Playing, Learning, and Enjoying with Mom | Promote skills to stimulate early learning through play | Games, quality time, and homemade toy creation | Responsive caregiving<br>Promote early learning | Good health<br>Responsive caregiving<br>Opportunities for early learning |
|  | 7 | Promotion of Healthy Nutrition and Health Care | Nutrition, Health, and Neurodevelopment | Guide on the importance of nutrition for child development and maternal well-being | Feeding practices, breastfeeding, and hygiene habits | Responsive caregiving<br>Integrated nutrition and responsive caregiving interventions | Good health<br>Adequate nutrition |
| Acknowledging Lived Experience, Emotions, and Resources | 8 | Emotional Regulation and Communication | The Sea of Emotions | Promote emotional awareness and communication skills | Emotional regulation, effective communication, and toxic stress management | Supporting maternal mental health | Good health<br>Security and safety |
|  | 9 | Social Support and Community Resources | Communities for Becoming Better People | Explore community resources as support in development | Support networks, virtual/territorial community, and family resources | Supporting maternal mental health | Good health<br>Security and safety |
|  | 10 | Healing: Breaking the Cycle of Adverse Childhood Experiences | Bonds of Love in Our Life Path | Address adverse experiences and break cycles of violence and trauma | Effects of toxic stress, gender violence, and adaptation | Supporting maternal mental health | Good health<br>Security and safety |
| Planning for the Future | 11 | Life Project: Toward Self-Sufficiency | Dream Catcher | Define personal and family goals for the future | Family planning, education, employment, and dream mapping | Supporting maternal mental health | Good health<br>Security and safety |
| Reflecting on Changes in Parenting | 12 | Looking Back on Course Content and Learning Process | Network of affectionate and responsive caregivers | Review of the learning experience and discussion about building a post-course support network | Strategies to continue strengthening the relationship with children | Responsive caregiving | Good health<br>Security and safety |

A multidisciplinary team comprising a family physician, two clinical psychologists, a neurocognitive scientist, and a medical anthropologist (all authors) designed the program in consultation with INAIPI staff and two advisors from the target population who had their first child as adolescents and whose children attended INAIPI centers. Sessions were delivered at an INAIPI center, consistent with WHO recommendations for group-based delivery to foster peer support and shared learning environments (WHO, 2024).

Course contents were informed by previous studies conducted with this population group: that adolescent mothers owned fewer developmental objects such as books and toys and preferred singing and technological resources (Castro & Sánchez-Vincitore, 2025; Nelson, Sánchez-Vincitore, et al., 2025); that the target population had been exposed to violence and ACEs, had difficulty recognizing the causes of affective symptoms, perceived a greater lack of emotional and family support, and was more exposed to verbal violence from partners (Nelson, Frías, et al., 2025); and that unplanned adolescent motherhood interrupted personal development plans and limited their ability to plan for the future, and that educational and material disadvantages persisted into adulthood even after adjustment for current age (Castro & Sánchez-Vincitore, 2025, 2026a). In response, the course includes the identification of useful household resources for play, the creation of songs as a pedagogical strategy, the promotion of reading as a tool for parenting and maternal development, a module to promote the identification and regulation of emotions and to help mothers recognize their own lived experience and how to heal, a module on support networks, relationship-building, and community resources, and a module on life projects to increase participants’ capacities, well-being, and goal setting.

### Materials

We collected pre- and post-intervention data using these instruments: a demographic survey used in previous studies in this community (Castro & Sánchez-Vincitore, 2025); the MICS Support for Learning module (UNICEF, 2019), measuring caregiver engagement in stimulation activities such as reading, storytelling, singing, and playing; the MICS Child Discipline Module (UNICEF, 2019), based on an adapted version of the Parent–Child Conflict Tactics Scale (Straus et al., 1998), with practices categorized into positive discipline (nonviolent methods) and violent discipline (psychological aggression and physical punishment); the Attitude Toward Spanking (ATS) 10-item scale (Holden et al., 1995); the Positive Parenting Scale (E2P), a 54-item self-report measure of parenting competencies across four subscales (Cronbach’s α = .95) (Gómez Muzzio & Muñoz Quinteros, 2014): bonding (secure attachment and socioemotional development), formative (learning, guidance, and positive discipline), protective (child safety, basic needs, and social support), and reflective (monitoring of parenting practices and self-care).

We also collected data using the ACE questionnaire, assessing 10 domains of childhood adversity through 19 dichotomous items (Felitti et al., 1998); the HITS intimate partner violence screening tool, assessing physical harm, insults and demeaning speech, threats of harm, and yelling or cursing (Cronbach’s α ≈ .80) (Sherin et al., 1998); the MICS module on attitudes toward domestic violence (UNICEF, 2019); and the Edinburgh Postnatal Depression Scale (EPDS), a 10-item screen for postnatal depression (Cronbach’s α = .87) (Cox et al., 1987).

Finally, we collected process indicators, including three satisfaction items rated on a 5-point Likert scale after each session, attendance records, and 24 binary session-level indicators (two per session, in most cases one knowledge and one skills indicator) to capture implementation fidelity and learning outcomes.

### Procedure

This feasibility study, conducted between September 12 and December 12, 2025, used a one-group pretest-posttest design to evaluate the 12-session intervention and assess its preliminary effectiveness in improving parental efficacy and parenting practices. On the first day, we provided a comprehensive explanation of the study protocol, obtained informed assent and consent, and collected baseline data using Audio Computer-Assisted Self-Interview (ACASI) via Tangerine, an open-source platform with offline functionality. The tablet application featured an interactive avatar that guided participants orally and visually through the survey, so that literacy was not a prerequisite for participation, while headphones ensured privacy during data collection. We collected post-intervention data using the same procedure. Three participants with lower session attendance completed their post-intervention assessments by telephone.

### Data analysis

We conducted paired comparisons between pre- and post-intervention scores across multiple outcome domains. Due to the small sample size (*n* = 11), we selected parametric (paired *t*-tests) or nonparametric (Wilcoxon signed-rank) tests based on the normality of change score distributions assessed via Shapiro-Wilk tests. We calculated effect sizes using Cohen’s *d* for paired samples. For binary outcome variables, we used McNemar’s test. We conducted all analyses in Python 3.12.3 using pandas 3.0.2 for data management, NumPy 2.4.4 for descriptive statistics and Cohen’s *d*, and SciPy 1.17.1 for statistical tests (shapiro for normality, ttest_rel for paired *t*-tests, wilcoxon for signed-rank tests, and binomtest for the exact McNemar test).

## Results

Twenty-seven mothers meeting the inclusion criteria were invited to enroll. Of these, 20 enrolled, 14 started the program, 3 were lost to follow-up after attending 1–2 sessions, and 11 completed both pre- and post-intervention assessments, yielding a retention rate of 78.6% among those who attended at least one session (11/14). Session attendance among completers averaged 8.64 sessions (*SD* = 2.66, median = 10, range: 4–12), representing 72.0% of the total intervention dose. One participant (9.1%) attended all 12 sessions, five (45.5%) attended 10–11 sessions (high dose, ≥83%), three (27.3%) attended 6–9 sessions (moderate dose, 50–75%), and two (18.2%) attended fewer than 6 sessions (low dose, <50%). Overall, 81.8% of completers (*n* = 9) attended at least half of the sessions. Session-level attendance ranged from 5 to 10 participants per session, with a mean of 7.92 (*SD* = 1.73).

### Participant characteristics

Participants had a mean age of 16.91 years (*SD* = 1.35, range: 14.75–19.97). All were primiparous, with study children averaging 1.15 years (*SD* = 0.84, range: 0.09–2.77). Five participants (45.5%) lived with the child’s biological father, one (9.1%) with a different partner, four (36.4%) with their own mother, and one (9.1%) lived alone with her child. Most had completed primary education only (63.6%, *n* = 7), while the remainder had completed secondary education (36.4%, *n* = 4). Participants reported limited household resources: all had access to a bed, and most had clothing (91%); a fan, gas tank, and kitchenware (82%); a stove and food (64%); a refrigerator or piped water (55%); fewer had furniture or a washing machine (36%). One participant (9.1%) owned her home, six (54.5%) rented, and four (36.4%) lived in a family home. The ages of the children’s biological fathers at the child’s birth averaged 21.00 years (*SD* = 7.20; *Mdn* = 19.00; range = 16.00–41.00). Among the fathers with education data (*n* = 6), 66.7% had completed primary education, and 33.3% had completed secondary education.

Participants reported high ACE exposure: 90.9% (*n* = 10) had at least one ACE, with a mean score of 3.18 (*SD* = 2.09, range: 0–7). Nearly half (45.5%, *n* = 5) met criteria for high ACE exposure (≥4 ACEs). The most prevalent ACEs were physical neglect (63.6%), emotional neglect (54.5%), verbal abuse (45.5%), physical abuse (45.5%), and family member incarceration (45.5%). The mean EPDS score was 8.64 (*SD* = 5.20), with 27.3% (*n* = 3) scoring at or above the clinical cutoff of 13 points. Five participants (45.5%) reported experiencing some form of intimate partner violence at baseline, and two (18.2%) at post-intervention.

### Participant satisfaction

The composite satisfaction score from three indicators (perceived usefulness, clarity of instruction, relevance of content) yielded an overall mean of *M* = 4.64 (*SD* = 0.48, *n* = 74 session-level responses). By session, composite scores ranged from *M* = 4.47 to *M* = 4.81, with all sessions maintaining means above 4.4 on the 5-point scale (Fig 1).

**Fig 1.**
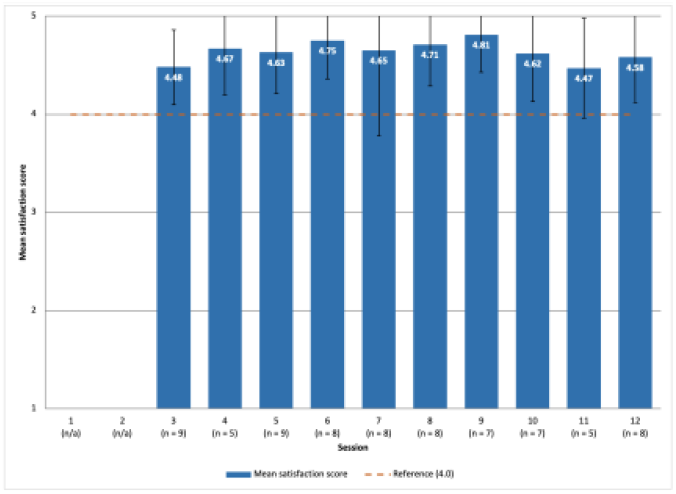
Mean satisfaction scores by session (*n* = 11)

### Session-based achievement indicators

We assessed implementation fidelity through session-specific learning indicators (Table 3). Knowledge indicators showed a higher mean achievement (64%) than skills indicators (45%), suggesting that participants more readily acquired conceptual understanding than applied competencies. Four sessions achieved 100% knowledge completion: session 4 (stress management), session 6 (play and early learning), session 9 (social support), and session 11 (life project). Session 11 was the only session to achieve 100% on both indicators, with all participants successfully creating a vision board that demonstrated life project concepts and long-term goals, along with associated activities. Sessions involving the creation of tangible products (homemade toys in session 6, vision boards in session 11, summary murals in session 12) achieved substantially higher success rates than those addressing abstract concepts. Session 7 (nutrition and health) showed the lowest overall achievement (25% for both knowledge and skills indicators), followed by session 8 (emotional regulation) with 25% knowledge and 38% skills. Sessions 2 (affective care) and 5 (positive discipline) achieved 33% on knowledge indicators.

**Table 3.** Implementation results indicators by session (*n* = 11)

| Session number | Topic | Type of indicator | Description | Participants completing the indicator |
| --- | --- | --- | --- | --- |
| 1 (n = 9) | Early Childhood Development (0-5 years) | Knowledge | The mother names 3 milestones of early childhood. | 78% |
|  |  | Skills | In a role-play, the mother successfully performs 2 activities that promote child development. | Activity not conducted |
| 2 (n = 9) | Affective Care | Knowledge | The mother names at least 5 children's needs. | 33% |
|  |  | Skills | In a role-play, the mother successfully performs 1 caregiving activity in an affectionate and responsive manner. | 44% |
| 3 (n = 10) | Basic Psychosocial Skills | Knowledge | The mother names at least 2 psychosocial skills. | 40% |
|  |  | Skills | In a role-play, the mother successfully performs 1 strategy to stimulate psychosocial skills. | 20% |
| 4 (n = 6) | Stress Management and Caregiver Self-Care | Knowledge | The mother identifies at least 3 factors that generate stress. | 100% |
|  |  | Skills | In a role-play, the mother successfully performs 1 practice aimed at reducing stress. | 67% |
| 5 (n = 9) |  | Knowledge | The mother identifies positive parenting techniques. | 33% |
|  | Positive Discipline in Early Childhood | Skills | The mother proposes positive parenting strategies in two hypothetical situations. | 44% |
| 6 (n = 8) | Play and Early Learning | Knowledge | The mother makes a homemade toy appropriate for her child's age. | 100% |
|  |  | Skills | The mother identifies 3 benefits of children's play. | 38% |
| 7 (n = 8) | Promotion of Healthy Nutrition and Health Care | Knowledge | The mother names 3 characteristics and 2 benefits of a healthy nutrition for the child. | 25% |
|  |  | Skills | In a role-play, the mother demonstrates 1 example of hygiene and care. | 25% |
| 8 (n = 8) | Emotional Regulation and Communication | Knowledge | The mother names 3 emotions and their functions. | 25% |
|  |  | Skills | In a role-play, the mother demonstrates 2 emotional self-regulation strategies. | 38% |
| 9 (n = 5) | Social Support and Community Resources | Knowledge | The mother identifies at least 1 community resource per domain (physical, social, services, governance, and socioeconomic). | 100% |
|  |  | Skills | The mother develops an individual map of the community resources that are available to her. | Activity not conducted |
| 10 (n = 8) | Healing: Breaking the Cycle of Adverse Childhood Experiences | Knowledge | The mother identifies at least 2 patterns of violence (or microviolence) within her family that she aims to change in her relationship with her child. | 75% |
|  |  | Skills | The mother names 5 supportive relationships she can rely on to foster positive emotional changes. | 25% |
| 11 (n = 5) | Life Project: Toward Self-Sufficiency | Knowledge | Through a vision board, the mother demonstrates an understanding of the concept of a life project and its application to her personal life. | 100% |
|  |  | Skills | The mother presents her vision board containing at least 1 long-term goal, 2 medium-term goals, and 2 activities for each goal needed to achieve them. | 100% |
| 12 (n = 10) | Looking Back on Course Content and Learning Process | Engagement | The mother actively participates in the creation and presentation of the murals. | 90% |
|  |  | Skills | The mother outlines a concrete action plan to establish a peer support network, including how members will communicate and at least one initiative with a defined start date. | 50% |

Note. n = number of participants attending the session. Percentages reflect the proportion of attendees who fully met each indicator criterion.

### Pre-post changes in primary outcomes

Table 4 presents pre-post comparisons for all outcome measures. Two outcomes showed statistically significant improvements: parental efficacy and cognitive stimulation. Parental efficacy, measured by the E2P scale, increased from *M* = 3.38 (*SD* = 0.44) to *M* = 3.84 (*SD* = 0.15), *t*(10) = 3.29, *p* = .008, *d* = 0.99. At the individual level, 81.8% of participants showed improvement, while 9.1% showed a decrease and 9.1% remained unchanged. At the subscale level, three of four domains showed statistically significant improvements with large effect sizes: protective competencies (*M* = 3.45 to 3.91, *p* = .003, *d* = 1.21), formative competencies (*M* = 3.27 to 3.95, *p* = .014, *d* = 0.90), and bonding competencies (*M* = 3.55 to 3.88, *p* = .023, *d* = 0.81). Cognitive stimulation (0–1 scale) increased from *M* = 0.42 (*SD* = 0.19) to *M* = 0.65 (*SD* = 0.22), Wilcoxon *p* = .036, *d* = 0.81. McNemar tests on binary outcomes showed no statistically significant changes: the proportion of children whom any caregiver read books to increased from 45.5% to 81.8% (*p* = .125), told stories from 36.4% to 81.8% (*p* = .062), and engaged in naming, counting, or drawing from 45.5% to 81.8% (*p* = .125); the proportion of mothers scoring at or above the EPDS clinical cutoff decreased from 27.3% to 18.2% (*p* = 1.000).

**Table 4.**
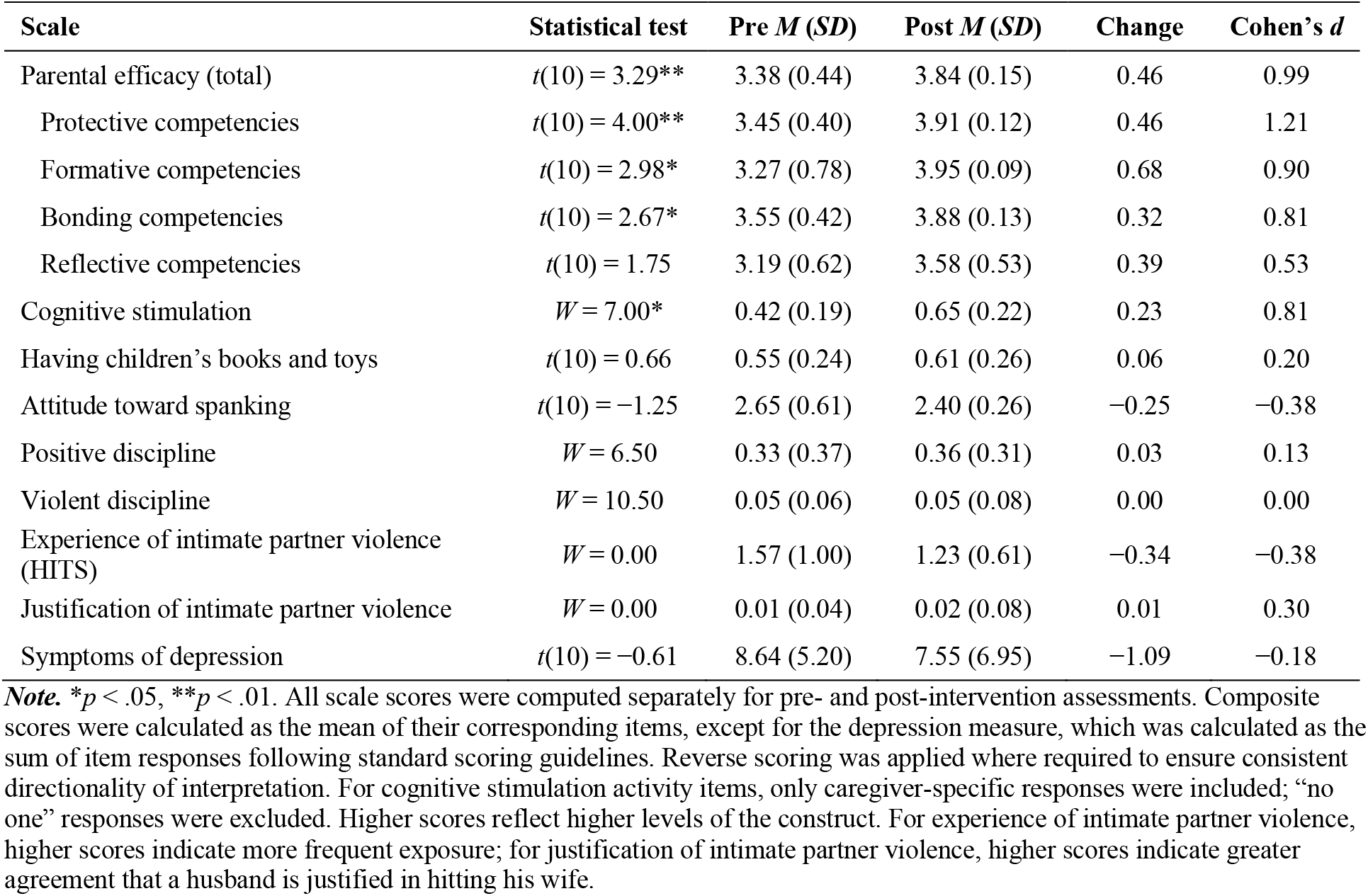
Pre-post intervention comparisons for primary outcomes (*n* = 11)

Home resources for ECD such as books and toys, attitudes toward spanking, positive and violent discipline practices, depressive symptoms, experience of intimate partner violence, and justification of intimate partner violence did not change significantly.

## Discussion

The ASCENDI intervention demonstrated feasibility and preliminary effectiveness, showing significant improvements in parental efficacy and cognitive stimulation scores from pre- to post-intervention, with large within-group effect sizes (*d* = 0.99 and 0.81, respectively). In addition, the program achieved high participant satisfaction (*M* = 4.64 on a 5-point scale) and a strong retention rate (78.6%), supporting its acceptability and implementation potential. These findings provide preliminary evidence supporting group-based nurturing care interventions tailored to adolescent mothers in resource-limited settings. Session-based learning indicators showed variable achievement, with knowledge indicators (25%–100%) generally outperforming skills indicators (20%–100%).

The improvement in parental efficacy (*d* = 0.99), which represents a large within-group effect, is practically meaningful. The fact that 81.8% of participants improved their efficacy supports the intervention’s consistent impact across individuals and reduces the concern that a small subset of participants drove the changes. Variance in parental efficacy scores also decreased substantially at post-intervention, indicating that participants converged toward higher levels of self-efficacy regardless of their baseline scores. This convergence aligns with catch-up effects documented in vulnerable populations (Zhang et al., 2021).

At the subscale level, the differential pattern of improvement across parental efficacy domains is informative. Protective competencies showed the largest effect (*d* = 1.21), followed by formative (*d* = 0.90) and bonding (*d* = 0.81) competencies, while reflective competencies did not reach statistical significance. Given the participants’ high exposure to adversity, the large effect observed in protective competencies may indicate that strengthening them is a key mechanism for interrupting intergenerational pathways of risk. Protective and formative competencies were directly targeted across multiple ASCENDI sessions addressing child development, stress management, positive discipline, and community resources. The non-significant change in reflective competencies is consistent with the broader finding that skills indicators were, on average, lower than knowledge indicators across sessions, as reflective competencies represent the most abstract and introspective domain of parenting and may require longer-term, sustained interventions to produce measurable effects, especially among adolescents exposed to poverty and adversity. This pattern suggests that a 12-session intervention can effectively strengthen concrete, action-oriented parenting competencies, while more abstract cognitive skills may benefit from extended or booster sessions.

The significant increase in cognitive stimulation activities represents a meaningful improvement in the home learning environment, particularly given prior findings that adolescent mothers in the Dominican Republic own few developmental resources and frequently rely on technological resources rather than books or active play (Castro & Sánchez-Vincitore, 2025; Nelson, Sánchez-Vincitore, et al., 2025). The ASCENDI curriculum directly addressed this gap by teaching mothers to identify household resources for play and promoting reading as a parenting tool. The pattern in session indicators—sessions involving tangible product creation outperforming those addressing abstract concepts—is consistent with learning transfer literature indicating that declarative knowledge precedes procedural skill development (Anderson, 1982). Sessions addressing nutrition, affective care, psychosocial skills, and emotional regulation may benefit from enhanced pedagogical approaches, potentially incorporating more concrete, product-based activities similar to those in higher-performing sessions. The generally lower performance on skills indicators compared with knowledge indicators suggests the need for additional scaffolding to support the transfer of conceptual learning to applied competencies.

The negligible changes in discipline practices reflect a floor effect at baseline, which may indicate social desirability bias in self-reported discipline behaviors, that mothers participating in INAIPI programs have already been exposed to messages about the harms of violent discipline for children, or that mothers who voluntarily enrolled were predisposed toward nonviolent discipline. It is also possible that the children’s young age limited the relevance of some of the more severe disciplinary behaviors assessed by the instrument. This is consistent with prior findings in this community that former adolescent mothers who participated in INAIPI programs used significantly less violent discipline than adult-onset mothers (*d* = −0.22) (Castro & Sánchez-Vincitore, 2026a). In national survey data from 15 Latin American and Caribbean countries, differences in violent discipline between adolescent and non-adolescent mothers were also non-significant in all countries with discipline data except Honduras after adjustment for socioeconomic covariates (Castro et al., 2026).

The heterogeneous response in depression symptoms (63.6% improved, 36.4% worsened) suggests that a 12-session parenting intervention may be insufficient to address the complex mental health needs of adolescent mothers with high baseline adversity (45.5% with ≥4 ACEs), and future iterations may benefit from integrating more intensive mental health components or referral pathways, consistent with evidence from this population that childhood adversity, rather than maternal age, is independently associated with intimate partner violence and depressive symptoms (Castro & Sánchez-Vincitore, 2026a).

Our effect sizes for parental efficacy and cognitive stimulation exceed those reported in a systematic review of 56 parenting intervention studies (pooled *d* = −0.46) (Backhaus et al., 2023), though direct comparison is limited by different outcome measures and our pre-post design without a control group. A Kenyan study found that combining messaging platforms with psychosocial support improved child development among adolescent mothers (Mwenda et al., 2023), and the Malaysian Naungan Kasih program reported comparable improvements in parental efficacy (Lachman et al., 2023), supporting cross-cultural applicability of these intervention components. Our findings extend this evidence by demonstrating that group-based interventions can improve both maternal self-efficacy and specific stimulation behaviors in a Latin American and Caribbean context. The evidence supporting nurturing care interventions for improving maternal health and child development outcomes (Fisher et al., 2023; McCoy et al., 2020; Tofail et al., 2023), combined with our results, suggests that such interventions hold promise for adolescent mothers across diverse settings.

This study has several strengths. First, the intervention was developed by a multidisciplinary team and reviewed by former adolescent mothers, enhancing cultural relevance and acceptability. Second, it incorporated topics identified in prior research in this community as particularly relevant to adolescent mothers in the Dominican Republic, including recognizing ACEs, emotional regulation, and life projects (Castro & Sánchez-Vincitore, 2025, 2026a; Nelson, Frías, et al., 2025; Nelson, Sánchez-Vincitore, et al., 2025). Third, implementation within the INAIPI infrastructure facilitates sustainability and integration with existing early childhood services, consistent with evidence that parenting interventions integrated with intersectoral programs yield better outcomes (Britto et al., 2017). Fourth, ACASI technology ensured participation regardless of literacy while maintaining privacy. Fifth, the group-based format aligns with WHO recommendations for fostering peer support and shared learning environments (WHO, 2024).

Several limitations should be considered when interpreting these findings. First, the small sample size (*n* = 11) limits statistical power and generalizability; effect sizes for outcomes showing medium effects may have reached statistical significance with a larger sample. Second, the one-group pretest-posttest design without a control group precludes causal attribution; observed improvements may reflect maturation, regression to the mean, or secular trends rather than intervention effects. Third, all outcomes were self-reported and thus susceptible to social desirability bias, particularly for sensitive topics such as discipline practices and intimate partner violence. Fourth, the study was conducted at a single site in Boca Chica, which may not be representative of other communities in the Dominican Republic. Fifth, the absence of long-term follow-up prevents assessment of whether improvements are sustained over time. Sixth, the intervention did not systematically include the children’s fathers or other caregivers, despite evidence that paternal involvement improves child development outcomes and maternal mental health, and that in this population the children’s fathers were often considerably older than the mothers and had lower educational attainment (Castro & Sánchez-Vincitore, 2025, 2026a; Chang et al., 2024; Kumar & Huang, 2021).

Future research should employ randomized controlled trials with larger samples, longer follow-up periods, direct assessment of child developmental outcomes, and exploration of the children’s fathers’ involvement. The father’s self-efficacy affects his involvement in play and parenting, making it relevant to encourage the paternal role and to provide support that facilitates the acquisition of parenting skills (Bowles et al., 2022; Yoo, 2022). The feasibility of implementing ASCENDI within INAIPI infrastructure suggests a scalable model for reaching adolescent mothers through established ECD programs, potentially interrupting the intergenerational transmission of adversity (Castro & Sánchez-Vincitore, 2026a; Nelson, Frías, et al., 2025), consistent with bioecocultural approaches that emphasize the intersection of developmental, cultural, and ecological factors in supporting adolescent mothers and their children (Wuermli et al., 2021).

## Conclusions

This feasibility study demonstrates that a 12-session group-based nurturing care intervention for adolescent mothers in the Dominican Republic can significantly improve parental efficacy and cognitive stimulation activities. Three of four parental efficacy subdomains showed large effect sizes, and high retention and satisfaction rates support program acceptability. Findings suggest that culturally adapted, WHO-based parenting programs delivered through existing early childhood services are feasible and preliminarily effective in resource-limited settings. Adolescent mothers represent an underserved population in parenting research; targeted, strengths-based interventions integrated within established infrastructure offer a scalable model for interrupting intergenerational cycles of adversity in low- and middle-income countries.

## Data Availability

We will upload the data to this repository once the manuscript is accepted for publication.

https://medrxiv.org

## Acknowledgments

The National Institute for Early Childhood Comprehensive Care of the Dominican Republic provided all logistical support for participant recruitment. We thank María Elena Valdez, Paulette Peterson, Cecilia Vallejo, María del Mar Camilo, Penélope Melo, Francina Guerrero, and María Teresa Mota from INAIPI for making this possible. We acknowledge José Navarro and Sara Menéndez for their feedback on an early version of the ASCENDI handbook, and Ana Elvira Montero and Anaísa Mercedes García for their contributions as consultants to the implementation of the ASCENDI feasibility study.

